# Process evaluation of the first universally designed asynchronous Multiple Mini Interview for optimised accessibility across neurotypes

**DOI:** 10.1101/2025.01.28.25321063

**Authors:** Alison Callwood, Jenny Harris, Madeleine Coe, Katie Sutton, Ivan Brewis, Lee Gillam, Paul Tiffin

## Abstract

Many jurisdictions legally require reasonable adjustments in personnel selection, to support individuals with disabilities, such as those with neurodevelopmental conditions. Recruitment to healthcare professions and education programmes is no exception. These measures often depend on applicants having formal diagnoses or a willingness to disclose their needs, overlooking the natural heterogeneity in cognition, learning, and behaviour. Consequently, traditional selection methods may inadvertently disadvantage certain candidates, Thus, more inclusive personnel selection practices are needed.

We aimed to assess the effect of co-designed interview modifications on differential performance between neurodivergent and neurotypical participants in a process evaluation. The co-design approach was employed to advance an existing online interview that utilised the Multiple Mini Interview (MMI) methodology in an asynchronous digital format. The interview was evaluated in two configurations: standard and modified. The modified version enhanced the standard version by incorporating a practice portal and accessibility features identified by neurodivergent volunteers. A total of 292 individuals, comprising 148 neurotypical and 146 participants self-identifying as neurodivergent from across the United Kingdom, took part in mock MMIs scored by independent assessors using a seven-point Likert scale.

Participants who self-identified as neurodivergent achieved significantly higher mean scores on the modified interview compared to the standard format (mean scores; 141.6 vs. 121.4 points; p<0.0001). In contrast, no statistically significant inter-group differences were observed for neurotypical participants for those taking the standard or modified interview. Scoring differences between neurodivergent and neurotypical participants reduced when the modified interview was used; no statistically significant intergroup difference in mean scores was observed in this condition (141.6 vs. 136.6; p=0.06). Inter-rater reliability for a random sample of double-blind scored interviews (10%) was high (ICC 0.8; p<0.001). Furthermore, 92% of neurodivergent participants reported that the optimised features facilitated the interview process, 70% perceived the outcomes as fair and objective, and 70% reported experiencing reduced anxiety compared to the unmodified interview.

These findings provide evidence that interview modifications can substantially reduce pre-existing disadvantages neurodivergent applicants may face when participating in online, digital interviews in this context. Such enhancements should be universally implemented to promote greater equity in personnel selection processes.

**Practitioner Points:**

1. Accessibility in online interviews can be optimised through low-cost high impact features including providing practise opportunities.
2. Optimising features should be available to all applicants to level the playing field regardless of ability/disability.
3. A co-design, universal approach can enable applicants to optimise their interview performance and should be integral to interview structure and set up.

## 1. INTRODUCTION

Achieving fair selection to health professions and education programs is a complex challenge. This is shaped by the interplay of unintended biases inherent in human assessment and the need to maintain workforce sustainability and suitability. A further complication is the recent, widespread adoption of online digital interviews, which often lack robust evidence to support their fairness, reliability and validity. Although equitable access to employment is recognised as a fundamental human right (United Nations, 2012), this ideal remains largely unattained for the neurodivergent community. Representing approximately 15-20% of the global population, 40% of neurodivergent individuals remain unemployed (Doyle, 2020). Furthermore, global data indicate that only 22% of people with autism are engaged in employment (Office for National Statistics, 2021). This underscores the critical need for more inclusive and evidence-based selection practices.

Neurodivergence is a broad term that refers to individuals whose brain functions differ from those of a ‘neurotypical’ person in everyday life (Clouder et al., 2020). It encompasses various conditions, including autistic spectrum conditions (ASC), dyslexia, dyspraxia, and attention deficit hyperactivity disorder (ADHD). Research suggests that neurodivergent individuals may be reluctant to disclose their support needs (McDowall et al., 2023). They may also ‘mask’ or ‘camouflage’ their behaviours due to fear of judgment (Livingston et al 2019). Feelings of potential disadvantage and negative perceptions when having to self-declare have been expressed (Doyle, 2023).

To accommodate neurodivergence, the provision of ‘reasonable adjustments’ is legally required in many jurisdictions. However, this often depends on applicants having a formal diagnosis. It also overlooks the natural heterogeneity in how people think, learn, and behave – acknowledging that everyone has a unique neurotype. Consequently, conventional personnel selection methods may inadvertently disadvantage neurodivergent candidates, highlighting the need for more inclusive practices.

Research by *Indeed* (2021) indicates that over 80% of interviews across sectors worldwide have shifted to an online format - a trend that is expected to continue (LinkedIn, 2024). However, this transition has taken place without understanding if, and how, individuals, particularly those in the neurodivergent community, can perform at their best when interviewed via this modality.

Little is known about the number of neurodivergent students studying medicine, nursing or allied health professions programmes. We do know that the number of students self-identifying as autistic more than doubled between 2014/15 and 2020/21 (HESA 2020). Medicine and dentistry have relatively low proportions of students self-identifying as disabled compared with most other disciplines, although this is increasing (HESA, 2020, Shaw 2022). The UK Medical Schools Council (MSC) recently highlighted a pervasive inequity problem regarding self-identified disability. They challenged UK medical schools to address ableism and to actively strive to make their environments, including recruitment, more inclusive (MSC, 2021).

In medical student selection, there is an approximate 50/50 split between online and face-to-face interviews (MSC, 2022). Although comprehensive data on healthcare admissions is lacking, research by Indeed (Indeed, 2021) indicates that over 80% of interviews have transitioned online across various sectors globally and are likely to continue in this format.

Online interviews can be conducted either synchronously or asynchronously. In a synchronous format, interviewers and applicants interact in real time using video conferencing technology. The asynchronous modality is a one-way interview where candidates record responses to pre-set questions at their convenience. These are then reviewed by interviewers later (Brenner et al., 2016).

Multiple Mini Interviews (MMIs) are a recent addition to online interviewing approaches (Selvam et al., 2021). MMIs are a structured interview format traditionally conducted face-to-face and originally developed for selection into medical programmes. They involve a series of short, timed interactions where candidates respond to various questions posed by different interviewers (Eva et al., 2004). This structured approach, along with a standardised scoring system, has been shown to reduce interviewer bias (Yusoff et al., 2019, Auton et al., 2024). MMIs have been widely adapted to online synchronous and asynchronous modalities since 2020. Concurrently, recent research has also demonstrated that asynchronous video interviews have the potential to mitigate interview bias (Brenner et al., 2016; Callwood et al., 2023).

Building on the evidential advantages of MMIs and the asynchronous modality, we sought to further understand how the performance of individuals with different neurotypes could be potentially optimised in a unique custom built asynchronous MMI. We propose that “optimisation” in this context involves two key elements: psychological safety and implementing user-interface adjustments to enhance accessibility.

Psychological safety refers to an environment where individuals feel secure and confident in bringing their authentic selves to the workplace (Edmonson et al., 2014). It consists of four key dimensions: *inclusion safety, learner safety, contributor safety, and challenger safety* (Newman et al., 2017). *Inclusion safety* ensures that everyone is valued and treated fairly. *Learner safety* allows individuals to ask questions, learn from mistakes, and explore new opportunities. *Contributor safety* fosters open dialogue and healthy debate, while *challenger safety* encourages people to speak up, share ideas, and suggest changes. We theorised that if applicants could familiarise themselves with the interview setup and user interface, it would promote inclusion and learner safety. This would ultimately make them feel more comfortable and better able to showcase their interpersonal skills and relevant knowledge evaluated by the interview.

User-interface adjustments can be facilitated using ‘accessibility toolbars’. These offer features designed to make technology more accommodating for individuals with different preferences and/or disabilities. These toolbars are increasingly common on websites and apps. Concerns have been raised regarding the efficacy of ‘quick fix’ solutions that depend on generic third-party applications. These may not always offer effective and inclusive support (Karlove, 2023). Additionally, these solutions often require users to self-identify their need for accessibility support-a step they maybe unprepared for, or unaware of. It is therefore essential to design tailored, person-centered solutions that proactively address accessibility needs. These should eliminate the sole reliance on users to identify and request the support they require.

## 2. METHODS

In a process evaluation, we aimed to assess the effect of co-designed interview modifications on differential performance between neurodivergent and neurotypical participants. Our hypothesis was that user-generated modifications could facilitate equitable performance among applicants with diverse neurotypes in online interviews, enabling them to demonstrate their full potential.

Our hypothesis was that user-created modifications could enable applicants with different neurotypes to perform to their full potential in online interviews.

We evaluated an existing asynchronous MMI platform in two configurations: standard and modified. In the modified version, we incorporated a practice portal and accessibility features identified through collaboration with neurodivergent volunteers.

Consenting participants were assigned anonymous usernames and passwords and a link to access the online MMI using Qualtrics (https://www.qualtrics.com).The MMI was set up in a three-question, four-minute circuit with a one-minute pause between questions. A requirement was that participants completed their answers to the three situational questions as if it were a real interview, taking care to answer the questions as fully as possible. All participants were provided with a short introductory video containing information about the interview format and upload process. They were prompted to check their video and microphones were functioning effectively. This was considered essential basic information required to use the asynchronous MMI platform successfully. Participant responses were automatically uploaded for later assessment by independent interviewers using a seven-point Likert scale from poor to excellent (1-7). Each applicant was scored against ten criteria with a total score range of 10-70 in each question and 210 across three questions. Interview assessors did not know which version of the platform participants had taken. The process evaluation was undertaken in five stages between July 2023 and December 2024 (Table 1).

**TABLE 1:** FIVE STAGES OF THE PROCESS EVALUATION.

| Stage | Date | Activity | Stakeholders |
| --- | --- | --- | --- |
| 1 | July - September 2023 | Evaluation of the standard asynchronous MMI | Neurodivergent participants |
| 2 | October 2023 | Focus group discussion |  |
| 3 | November - August 2024 | Practice portal refinement, toolbar design and build | Software Developers |
| 4 | September - November 2024 | Evaluation of the asynchronous MMI with the toolbar built in (modified version) | Neurodivergent participants |
| 5 | December 2024 | Evaluation of effectiveness of the modified version | Neurodivergent and neurotypical participants |

### Data collection

#### Stage 1

Consenting neurodivergent participants completed the standard version of the interview with no practice portal or accessibility optimisations. Feedback was captured using a follow-up questionnaire which incorporated a range of questions relating to interview experience and utilised a five-point Likert scale ranging from ‘Strongly Disagree’ to ‘Strongly Agree’. Participants were asked to rate a range of potential accessibility features that they felt could be added to the interview to optimise their performance using a five-point Likert scale (‘Not very helpful’ to ‘Very helpful’). The potential accessibility features were derived from volunteers with lived experience of neurodivergence from the project Advisory Board. The questionnaire also included free-text boxes for feedback and suggestions on additional features.

#### Stage 2

All neurodivergent participants from Stage 1 were invited to take part in a focus group facilitated by an experienced researcher. The aim was to augment the feedback from Stage 1 and achieve information power (Braun and Clarke, 2019). Stage 1 data were used to inform the topic guide.

#### Stage 3

Practice opportunities and accessibility optimisations were built in including the toolbar featuring the most requested optimisations thereby creating the ‘modified’ version.

#### Stage 4

A separate group of neurodivergent participants completed the same three-question, four-minute MMI circuit as Stage 1 on the ‘modified’ version, followed by an evaluation questionnaire.

#### Stage 5

Neurotypical participants were randomly allocated to complete the standard version and modified versions using the same three-question, four-minute MMI circuit. Inter-rater reliability was assessed from a random sample of interviews, rated blindly by two separate raters.

### Design

The co-design approach (Robert et al., 2022) used was rooted in Participatory Action Research (PAR) (Schneider et al., 2012). This aimed to facilitate meaningful user engagement in developing the interview format and modifications. Service providers and users, including neurodivergent volunteers, collaborated on all aspects of the study development, design, and implementation. This collaboration fostered a human-centred understanding of key stakeholder experiences, guiding technological improvements to enhance accessibility. Co-design, particularly when integrated with participatory methodologies like PAR, has been shown to improve service quality and create more satisfactory services (Benz et al., 2024). By involving diverse stakeholders in the design process, we ensured that the interview addressed the specific needs and preferences of its intended users.

Principles of Universal Design for Learning (UDL) (Rose & Meyer, 2002) informed this research, with the vision that optimisation features should be available to all applicants by default. This would enable those applicants who might normally ‘mask’ their needs or be reluctant to disclose neurodivergence to access the accessibility features by default.

The study was grounded in Gilliland’s justice-based model (Gilliland, 1993), which highlights the importance of adhering to both procedural justice rules (the characteristics of the selection process) and distributive justice rules (ensuring equity and equality) to enhance applicants’ perception of the selection system.

### Participant recruitment

All participants were identified through the platform Prolific (www.prolific.com). Prolific is a commercial company that facilitates the signing up of pools of research participants across the United Kingdom (UK). In accordance with the Prolific’s terms and conditions, we outlined our requirements through uploading a recruitment letter containing links to further information and a consent form. The platform facilitates the application of inclusion and exclusion criteria, including self-identified neurodivergence. This enabled the sourcing of the required participant samples. Participants were remunerated according to Prolific’s terms and conditions policy at £15 per hour pro rata. Inclusion criteria included being able to speak, read and understand English; have access to a desktop or laptop, and self-identify as neurotypical or neurodivergent (including but not limited to: ADHD, ASC, dyslexia and dyspraxia). Exclusion criteria applied to Stage 4 where we restricted participation to volunteers who had not taken part in Stage 1.

### Analysis

Evaluation data were analysed using descriptive statistics and conventional content analysis (Hsieh and Shannon, 2005) to gain a richer understanding of the perceptions and experiences of participants. The free-text data collected within the focus group (Stage 2) was subject to thematic analysis (Braun and Clarke, 2006). Two members of the research team independently scrutinised identified themes prior to revising and agreeing the finalised themes collaboratively. Results from Stage 1 and the focus group were collated and shared with the interview platform software developers, who were then able to build the modified version.

We used t-testing to formally test for inter-group differences in interview scores between neurotypical and neurodivergent participants, with and without the accessibility features. Effect sizes were calculated using Cohen’s d. Inter-rater reliability was assessed via calculation of an intraclass correlation coefficient (ICC). All analyses were conducted using Stata V.17 (StataCorp.2021). In terms of study power, it was estimated a minimum sample size of 46 in each group would provide 80% power to detect an effect size of 0.6 with a significance level of 0.05 for two-tailed testing (Das et al., 2016).

### Patient and Public Involvement and Engagement

Patient and Public Involvement and Engagement (PPIE) was an integral component of our co-design study. A project Advisory Board was convened at the start of the research comprising volunteers with lived experience of neurodivergence alongside neurotypical volunteers with interest and experience in recruitment. The Advisory Board met regularly throughout the duration of the project, actively contributing to the study design including the development of the information sheet, questionnaire and MMI questions.

## 3. RESULTS

### Sample characteristics

Data were available from n=146 participants who self-identified as neurodivergent (n=48 in Stage 1, n=98 in Stage 4) (Table 2). Participants reported neurodivergent challenges including but not limited to ASC, ADHD and dyslexia. Five participants volunteered to attend the focus group in Stage 2. N=148 neurotypical participants took part in Stage 5.

**Table 2:**
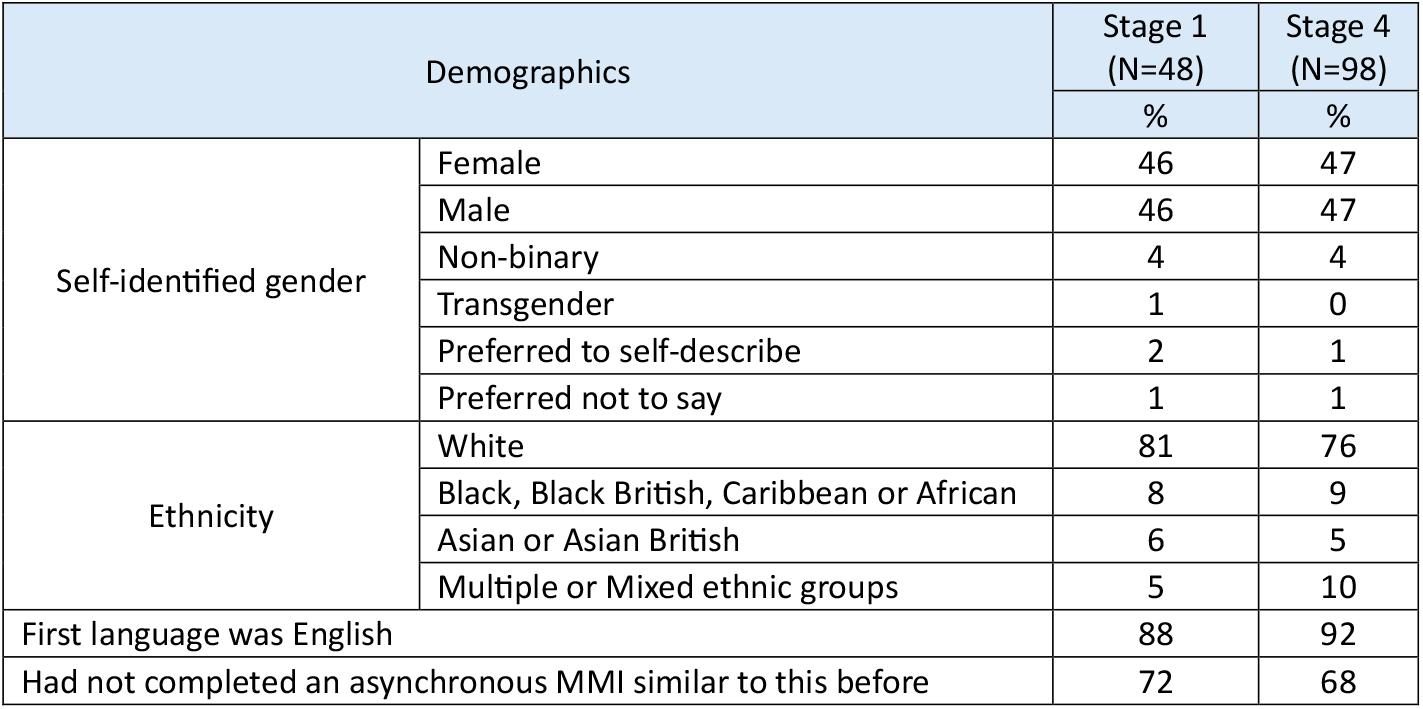
Participant self-identified characteristics.

In Stage 1, the majority (90%) felt the instructions were easy to follow, and that the platform facilitated easy completion of the interview (81%) (Table 3). Many participants (63%) reported feeling less anxious whilst completing the online, asynchronous MMI compared to other interview techniques. Additionally, participants (65%) felt the interview format would lend itself to promoting fairer, more objective interview outcomes than traditional interviewing techniques. Participants found existing features of the platform beneficial such as the short breaks (73%) and provided feedback about other potentially helpful optimisations that could be integrated into the interview platform such as real-time sub-titles, modifying how they saw themselves in the video and being able to choose font, background colour/contrast.

**Table 3.** Participant evaluation of the interview platform.

| Overall | Response | Stage 1 (%) | Stage 4 (%) |
| --- | --- | --- | --- |
| Felt the instructions were easy to follow | Agree-Strongly agree | 90 | 93 |
| Felt the platform made it easy to complete the interview |  | 81 | 92 |
| Felt less anxious doing interviews on this platform than doing Zoom/<br>other videoconference facilitated interviews |  | 63 | 67 |
| Felt less anxious doing interviews on this platform than doing face to<br>face interviews |  | 62 | 69 |
| Felt platform would make interview outcomes fairer and more<br>objective |  | 65 | 70 |
| **Top three suggested helpful adjustments (Stage 1)** | **Response** | **%** |  |
| Option for real-time subtitles | Yes | 64 | - |
| Being able to modify the video container | Helpful-<br>very helpful | 61 | - |
| Being able to change the font/brightness/contrast |  | 51 | - |
| **How helpful being able to change accessible optimisations (Stage 4)** | **Response** | **%** |  |
| Font style and size |  | - | 65 |
| Subtitles addition | Helpful-<br>very helpful | - | 59 |
| Minimisation of video container |  | - | 44 |
| Background blur |  | - | 37 |
| Background colour and contrast |  | - | 23 |

### Stage 2: Focus Group

The themes identified in the focus group are shown in Figure 1:

**Figure 1:**
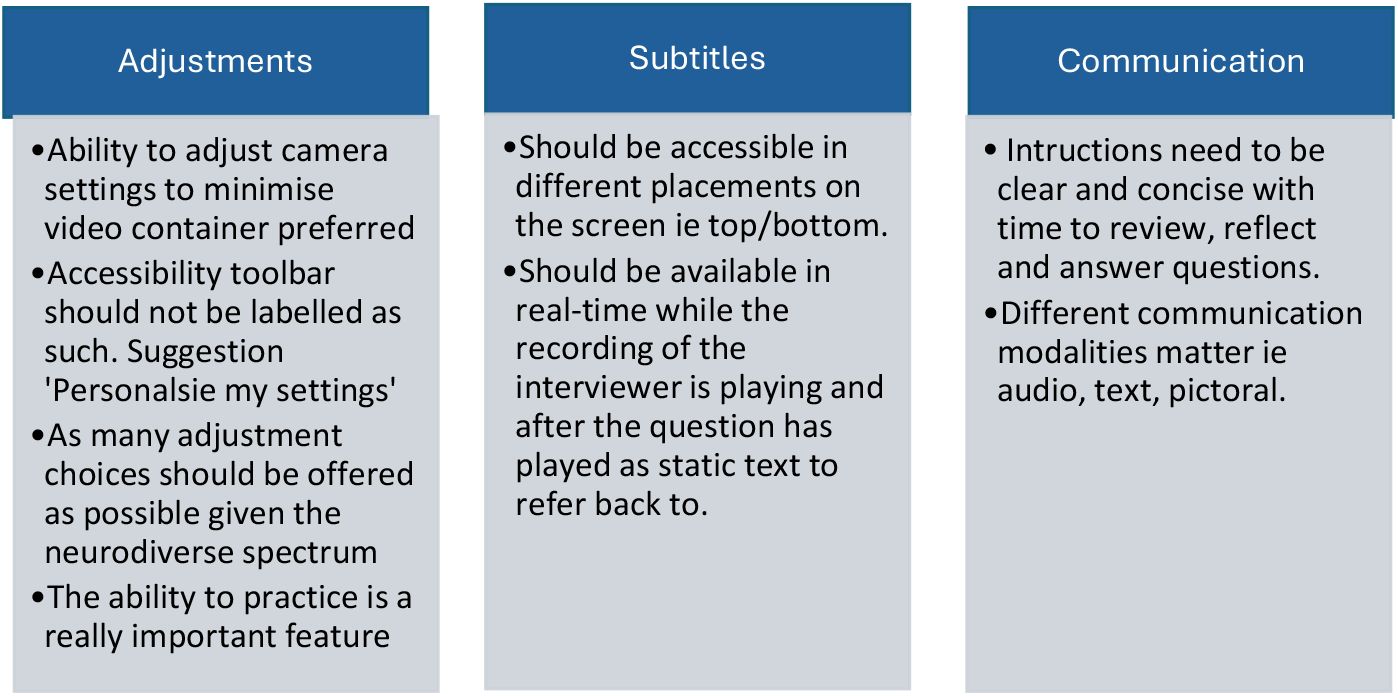
Focus group theme findings.

The themes from the focus group were used to augment the Stage 1 questionnaire findings and helped to identify priority optimising features.

### Stage 3

A practice portal was built to facilitate familiarisation with the interview process, allowing unlimited access to support this goal. It included features such as a microphone and video check, practice question and answer upload, and a toolbar that delivered optimisation tools previously identified by neurodivergent participants. (Figure 2). The toolbar was labelled ‘Personalise my settings’ following feedback from both the Focus Group and the Advisory Board members. It was suggested that this might encourage more applicants to check the options out to see if any meet their preferences thereby embedding the universal design approach.

**Figure 2:**
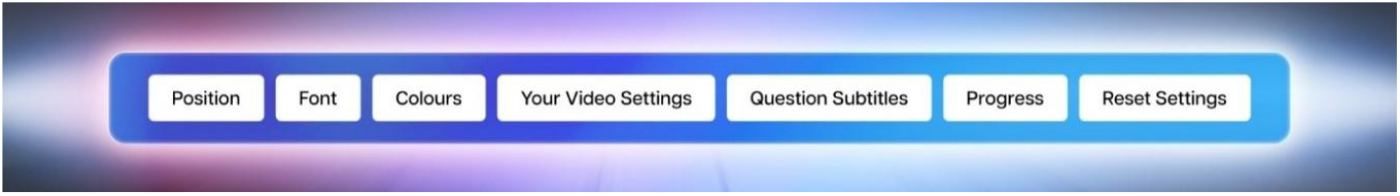
Toolbar features

- **Position:** refers to the position of the toolbar on the screen i.e. top/bottom/vertical/ horizontal.
- **Font:** Six different font choices are offered, including Bionic Reading and Dyslexic font in a choice of sizes
- **Colour:** Eight colour font/background pairings were incorporated reflecting volunteer preferences for colour/contrast.
- **Your Video Settings**: Enables minimisation of the video container for those who did not like seeing themselves fully on the screen and the option to blur their background.
- **Question Subtitles**: Subtitles were made available in real time and as reference text after the pre-recorded MMI questions were played.
- **Progress**: Offers choice as to how the timing of the MMI questions are displayed. To avoid time-blindness, visualisation of progress within the timed circuit was necessary. However, a count-down timer was perceived by many neurodivergent participants as stressful. We devised the option to be able to flip from ‘count down’ to ‘count up’ to alleviate this.
- **Reset Settings:** Revert to original settings in case selections made are less desirable on reflection.

### Stage 4

In comparison to the Stage 1, the data collected during Stage 4 showed an increase in participants feeling that the platform made it easy to complete the interview (92% versus 81%) and more participants reported feeling less anxious than at face-to-face interviews (69% versus 62%). Further, the data highlighted which optimisations participants found particularly helpful in increasing accessibility for neurodivergent individuals; font style and size (65%), the ability to add subtitles (59%), to select video options (44%) were among the most frequently cited (Table 4).

**Table 4:** Stakeholder free text evaluation (Stage Four)

| Themes | N=314<br>(83%) | % of total<br>comments | Illustrative quotes |
| --- | --- | --- | --- |
| Positives |  |  |  |
| Online, automated aspect and associated personal autonomy | 79 | 21 | *"Not having to interact with someone in real time meant I could focus more on my answers than trying to 'read' the other person. Can be done from the comfort of home (this makes a big difference to my anxiety symptoms)."*<br>*"The abstract structure of the interview is different. I am not sitting in front of a panel being questioned, rather, I am in control of an interview where I am in the centre reading questions and providing answers".* |
| User interface, usability of platform | 75 | 20 | *"Very easy to use platform with clear instructions."*<br>*"The functions and tools provided worked in a way that was easy for a first-time user. The interface was clutter free and intuitive".* |
| Subtitles and question transcript | 56 | 15 | *"I found the subtitles very useful to better understand the questions and to be able to read them through while answering, this is something that doesn't happen in other type of interviews."*<br>*"I found having the text of the question after the video of the person asking it super helpful".* |
| Introduction video | 26 | 8 | *"I liked the fact that the instructions were explained both by video and text as it makes it accessible to more people."* |
| Communication | 22 | 7 | *"The interviewers were friendly and spoke clearly, this made me feel less nervous."*<br>*"The different people asking the questions kept me engaged."* |
| Breaks | 21 | 6 | *"Having a break between questions without the interviewer seeing you is good; it means you shake off negative feelings from a previous answer."* |
| Timer/prompts | 20 | 5 | *"The timer helped me keep an idea of the time and reduced time-blindness issues."* |
| Practice question | 8 | 2 | *"I found the initial practice interview and going over the various controls helpful the UX was intuitive and engaging."*<br>*"The feeling of control - being able to start when you're ready and practice first."* |
| Video settings | 7 | 1 | *"I liked that my web camera box showed on screen so I could see that I was placed correctly/etc."* |
| Suggestions for improvement | N=63<br>(17%) | % of total<br>comments | Illustrative quotes |
| Make accessibility toolbar more obvious | 34 | 9 | *"Minimising videos, counting up or down, changing font size or colour ... etc. nothing of that was clear to me. Perhaps it would be useful if there was a hint highlighter that takes me through the interface at the beginning to highlight the features!"* |
| Shorter introduction and questions | 23 | 6 | *"The instruction video could have been broken down to parts, it was a bit too long to follow in one go."* |
| Subtitles accuracy | 6 | 2 | *"Some of the transcript wording did not match the subtitles, that led to some confusion".* |

Within optional free-text boxes, a total of 377 comments were collected about participants’ experiences of the modified platform.

### Stage 5

Following completion of the MMI by neurotypical participants in both the standard and modified versions, interview scores were analysed. The results are shown in Table 5. Neurodivergent participants obtained statistically significantly higher mean scores on the modified interview (mean 141.6 vs 121.4, p<0.0001) signalling a 15% increase in attainment. The effect size, indicated by a Cohen’s d value of 0.7 indicated a moderate to large effect. No statistically significant differences were observed between interview formats for neurotypical participant (group mean score; 136.6 vs 133.1 p=.54). Non-statistically significant differences in mean interview scores were observed between the neurodivergent and neurotypical groups with optimisations built in (mean 141.6 vs 133.2, t (134) =-1.91 p>0.06, Cohen’s d -.29 indicates small effect size). Inter-rater reliability of a random sample of interviews (10%) across the two versions was very good (ICC 0.8 p < 0.001, 95% CI .59-.87).

**Table 5.**
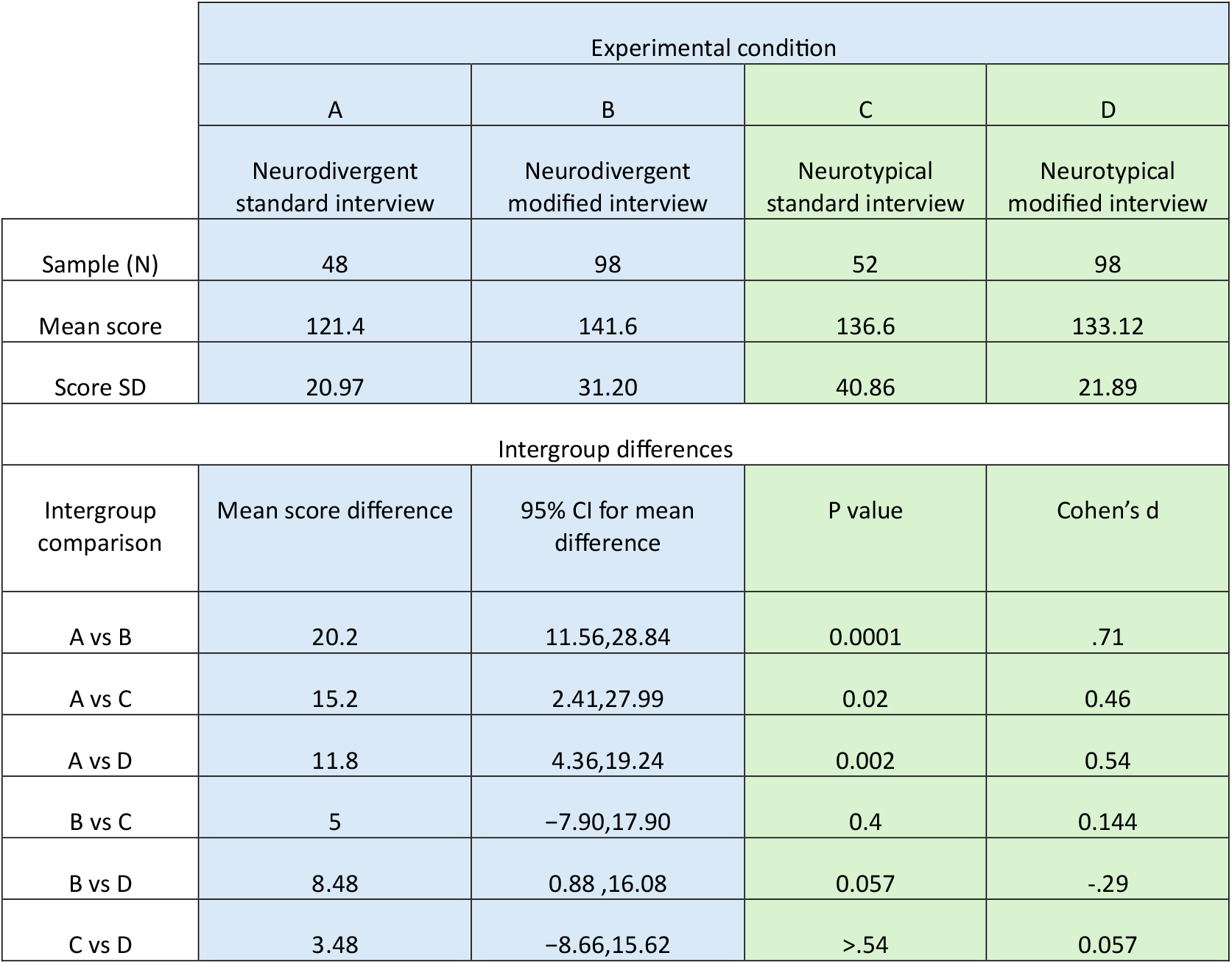
Online interview performance and intergroup difference for the four experimental conditions.

## Discussion

Our aim was to explore if and how online interviews could be optimised to reduce differential performance between neurotypical and neurodivergent individuals. Our approach was grounded in an understanding of neurodiversity as a trait, that is shared, to some degree, by all potential candidates. Central to this effort was a co-design approach, where we avoided assumptions about what might be effective and instead consulted individuals with relevant lived experience.

Statistically significant differences were found between neurodivergent participants mean scores with and without the optimising features. This indicated that the accessibility optimisations enhanced interview performance in this group of individuals. The lack of statistically significant differences between neurotypical participants and neurodivergent participants who had access to the accessibility features suggests that these features levelled the playing field in this context.

There is limited research on how neurodivergent candidates experience accessing jobs and education programs, however our findings build upon the recommendations of Tomlinson et al. (2024). Participants in this study reported that the optimisations made the interview process easier and reduced their anxiety. They also felt that the platform would make interview outcomes fairer and more objective. This is a particularly important finding given published evidence that applicants who felt that the selection process was fair are more likely to view an organisation favourably and accept recruitment offers (Truxillo et al., 2002). Optimisation options that were reported to be particularly beneficial to neurodivergent participants were the real-time subtitles and ability to minimise how the saw themselves. This corroborates previous research findings from Yurucki, et al., (2023) where the addition of subtitles allowing participants to read as well as hear the questions were positively evaluated.

It should be noted, that whilst the responses to the enhanced asynchronous online MMI platform were largely positive, there was some negative feedback. Firstly, and consistent with other online interview modalities (Yuruki et al., 2023), participants reported that they found seeing themselves on screen off-putting. This was addressed through the ability to minimise themselves. The countdown process of the MMI itself was stress-inducing for some but this was successfully mitigated by the option to ‘count up’.

Our vision was to enable as many applicants as possible to achieve their full potential, mindful that many may or may not be aware of, or willing to share their neurotype. Our results did show a dispersion in preferences for the different optimisation options, which could be indicative of the varied accessibility requirements of different neurodivergent presentations. It also signals the potential benefits of universal design where the toolbar and practice portal were designed to be used by all applicants, not just those who self-identified as neurodivergent. Implementing this universal design and calling the toolbar ‘personalise my settings’ rather than ‘accessibility toolbar’ avoids the need to self-declare any neurodivergent conditions, which may induce feelings of potential disadvantage and fear of being judged (Doyle, 2023). This approach encourages psychological safety and normalises individual differences.

We were mindful that we did not want to inadvertently advantage neurotypical applicants because of the available accessibility optimisations thereby potentially widening the attainment gap. This was not borne out by our data-indeed neurotypical applicants had slightly lower, non-statistically significant mean scores differences between the interview conditions. This signals the potential of universal design to narrow the attainment gap between neuro types grounded in the assumption that we are all neurodiverse (Doyle, 2023). As a result, we recommend incorporating these principles into policy and practice as mandatory features. Further longitudinal research studies are recommended to track the in-role or in-program performance of candidates, ensuring that individuals with diverse needs are adequately supported to reach their full potential in educational or professional settings once recruited.

### Potential strengths and limitation

We acknowledge the potential limitations of a three-question MMI set up in relation to MMI theory which demonstrates enhanced psychometric properties with six or more questions. (Eva et al., 2004). This was a pragmatic decision since we were not exploring reliability or validity but need to secure participants to the study and did not wish to over-burden them.

Participants were recruited through the online platform Prolific, which may have introduced selection bias, as they were potentially more comfortable interacting online. Self-identification was required, though this largely aligns with current real-world practice in relation to accommodations in selection processes, made via self-declarations.

Lastly, our study power was only calibrated to detect moderate (Cohen’s d≥0.6) intergroup differences with acceptable precision. Thus, smaller differences would not be detected at statistically significant levels, should they have existed. However, more minor intergroups differences are less likely to be substantively meaningful in a personnel selection context.

### Rigor

Enhancing the psychological safety of our neurodivergent participants was paramount throughout this study. During the focus group, careful consideration was taken to ensure that participants felt comfortable and secure. This included incorporating rest breaks, agreeing ground rules in both verbal and written format at the start and allowing participants to choose to have their camera on or off as they preferred (Le Cunff et al., 2023).

A requirement of this study was that participants completed the mock interviews as if they were a live situation. Participants’ interviews were screened by an independent third-party researcher experienced in MMI assessment prior to being marked to ensure these criteria were met. Assessors did not know which version they were marking to avoid unintended bias. Furthermore, ten percent of the interviews were second marked by an independent third-party assessor to evaluate inter-rater reliability.

## Conclusion

We have a collective responsibility to ensure that our workforce represents the diversity of the society it serves. As selection to many jobs and education programmes appear to be continuing to use an online format, it is essential that interview processes are developed and implemented in a way that supports optimal performance for all applicants regardless of ability or disability.

These findings signal that our asynchronous digital interview, grounded in MMI methodology and integrating practice opportunities and accessibility features, positively influence the interview performance of neurodivergent individuals in this context. Furthermore, the modifications resulted in smaller score differences between neurotypical and neurodivergent participants. These results highlight the potential of the modified interview to mitigate the relative disadvantage experienced by neurodivergent individuals in online interviews, thereby fostering greater equity in personnel selection processes.

## Data Availability

All data produced in the present study are available upon reasonable request to the authors

## Acknowledgments

Grateful thanks to the volunteers and professional colleagues without whom this work would not be possible.

